# Carboxypeptidase B2 gene polymorphisms in the donor associate with kidney allograft loss

**DOI:** 10.1101/2023.05.08.23289675

**Authors:** Felix Poppelaars, Siawosh K. Eskandari, Jeffrey Damman, Ashley Frazer-Abel, V. Michael Holers, Bradley P. Dixon, Mohamed R. Daha, Jan-Stephan F. Sanders, Marc A. Seelen, Bernardo Faria, Mariana Gaya da Costa, Joshua M. Thurman

**Affiliations:** Department of Internal Medicine, Division of Nephrology, University Medical Center Groningen, University of Groningen, Groningen, The Netherlands; Department of Medicine, University of Colorado School of Medicine, Aurora, Colorado, USA; Department of Pathology, Erasmus Medical Center, University Medical Center, Rotterdam, The Netherlands; Exsera BioLabs, University of Colorado School of Medicine, Aurora, Colorado, USA; Departments of Pediatrics & Medicine, University of Colorado School of Medicine, Aurora, Colorado, USA; Department of Nephrology, University of Leiden, Leiden University Medical Center, Leiden, The Netherlands; Nephrology and Infectious Disease R&D Group, INEB, Institute of Investigation and Innovation in Health (i3S), University of Porto, Porto, Portugal; Department of Anesthesiology, University of Groningen, University Medical Center Groningen, Groningen, The Netherlands

**Keywords:** carboxypeptidase, complement, transplantation, nephrology, genetics.

## Abstract

**Introduction:** Plasma carboxypeptidase B2 (CPB2) is an enzyme that cleaves C-terminal amino acids from proteins, thereby regulating their activities. CPB2 has anti-inflammatory and anti-fibrinolytic properties and can therefore be protective or harmful in disease. We explored the impact of functional carboxypeptidase B2 gene (*CPB2*) polymorphisms on graft survival following kidney transplantation.

**Methods:** We performed a longitudinal cohort study to evaluate the association of functional *CPB2* polymorphisms (rs2146881, rs3742264, rs1926447, rs3818477) and complement polymorphisms (rs2230199, rs17611) with long-term allograft survival in 1,271 kidney transplant pairs from the University Medical Center Groningen in The Netherlands.

**Results:** The high-producing *CPB2* rs3742264 polymorphism in the donor was associated with a reduced risk of graft loss following kidney transplantation (hazard ratio, 0.71 for the A-allele; 95%-CI, 0.55–0.93; *P*=0.014). In fully adjusted models, the association between the CPB2 polymorphism in the donor and graft loss remained significant. The protective effect of the high-producing *CPB2* variant in the donor could be mitigated by the hazardous effect of gain-of-function complement polymorphisms. Additionally, we compiled a genetic risk score of the four *CPB2* variants in the recipients and donors, which was independently associated with long-term allograft survival. Furthermore, this genetic risk score substantially improved risk prediction for graft loss beyond currently used clinical predictors.

**Conclusion:** Kidney allografts from deceased donors possessing a high-producing CPB2 polymorphism are at a lower risk of graft loss after kidney transplantation. Furthermore, our findings suggest that CPB2 might have a protective effect on graft loss through its ability to inactivate complement anaphylatoxins.

**Essentials:**

- Carboxypeptidase B2 (CPB2) is a metalloprotease with anti-fibrinolytic and anti-inflammatory properties.
- We investigated the impact of *CPB2* polymorphisms on graft loss after kidney transplantation.
- The rs3742264-A SNP in the donor, linked to higher CPB2 levels, decreased the risk of graft loss.
- CPB2 could have a protective effect on graft survival by inactivating complement anaphylatoxins.

## Introduction

Transplantation is the best treatment option for patients with kidney failure due to its association with reduced mortality, the risk attenuation of cardiovascular disease and improvement of quality of life.[1–4] In recent decades, surgical advances and improved immunosuppressive regimens have drastically lengthened graft survival from months to years, whereas the long-term outcomes after kidney transplantation have not improved at the same rate.[5] A critical driver of poor long-term outcomes is the damage incurred by the kidney allografts prior to, during, and after transplantation as a consequence of donor conditions (i.e., brain death), ischemia-reperfusion, and allosensitization. A growing body of evidence implicates the innate immune system as well as the coagulation cascades are a major driver of graft injury, since they are independently associated with long-term transplant outcome.[6,7] Notably, the cross-talk between these two systems is increasingly recognized as an vital contributor to graft loss and is, therefore, seen as a potential therapeutic target in transplantation.[8]

As enzymes with both anti-fibrinolytic and anti-inflammatory qualities, carboxypeptidases are a target of interest within this scope. Carboxypeptidases are a family of zinc-containing proteolytic enzymes that cleave carboxy-terminus (C-terminus) amino acids from biologically active proteins, thereby regulating their activities.[9] These enzymes can be soluble or membrane-bound and have a preference for either hydrophobic or basic amino acids.[9,10] Carboxypeptidase B2 (CPB2), also known as thrombin-activatable fibrinolysis inhibitor (TAFI), is synthesized by the liver and is one of only two carboxypeptidases present in plasma (Fig. 1A).[10] CPB2 circulates as an inactive proenzyme (proCPB2) and is predominantly activated by plasmin bound to glycosaminoglycans or by the thrombin–thrombomodulin complex (Fig. 1B).[11] Since there are no known physiological inhibitors, CPB2 activity is assumed to be regulated by its very short half-life of 8 – 15 minutes at 37 °C.[12] Once activated, CPB2 inhibits fibrinolysis by cleaving C-terminal lysine residues from partially degraded fibrin, thereby preventing the binding of fibrinolytic components (Fig. 1C – D).[13–16] Besides the anti-fibrinolytic activity during fibrin clot formation, CPB2 can also exert anti-inflammatory properties.[10,17–20] More specifically, a number of studies have demonstrated that complement anaphylatoxins C3a and C5a are physiological substrates of CPB2.[10] CPB2 cleaves the C-terminal arginine residue of C3a and C5a, generating the metabolites C3adesArg and C5adesArg and limiting the pro-inflammatory effects of these anaphylatoxins (Fig. 1C and E).[10]

**Figure 1:**
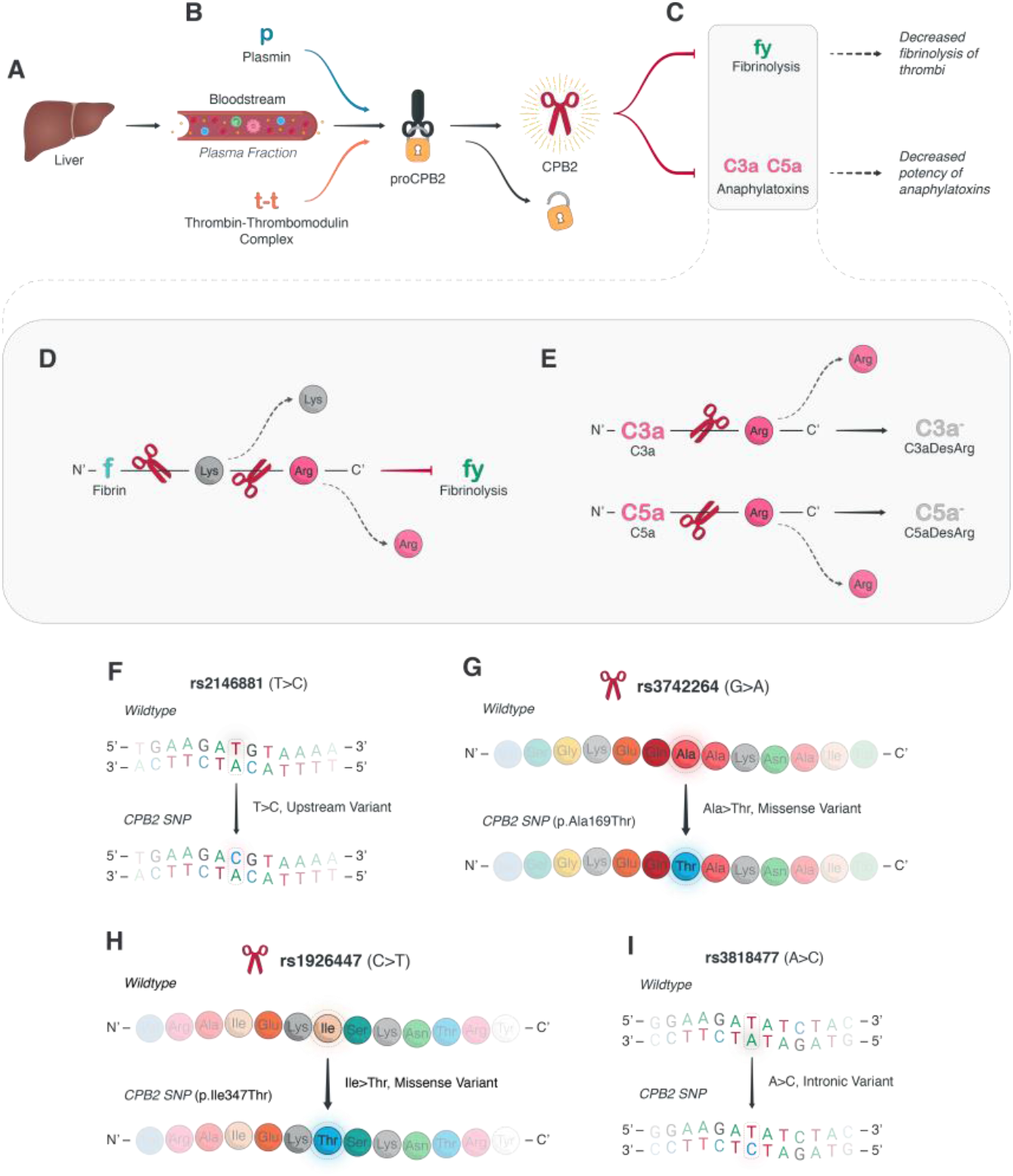
Illustration of the CPB2 pathway and examined *CPB2*-related polymorphisms. (**A**) A precursor form of CPB2 (proCPB2) is produced in the liver and released into the plasma fraction of the blood. (**B**) There, it can interact with plasmin and the thrombin-thrombomodulin complex to gain its enzymatic, carboxypeptidase activity as mature CPB2. (**C** – **E**) The main effects of CPB2 are exerted on (**C**) fibrinolysis and anaphylatoxin. (**D**) The effect on fibrinolysis is mediated through fibrin by CPB2-dependent removal of C-terminal lysine and arginine residues, thus thrombus resorption by fibrinolytic mechanisms. (**E**) The neutralization of anaphylatoxins such as C3a and C5a is mediated by C-terminal lysis of arginine residues, creating C3aDesArg and C5aDesArg variants with decreased inflammatory potency, resulting in anti-inflammatory effects. (**F** – **I**) In this study, we assessed the associations of four single-nucleotide polymorphisms (SNPs) in kidney allograft donors and recipients on graft survival. These SNPs were (**F**) rs2146881 (G>A), (**G**) rs3742264 (p.Ala169Thr), (**H**) rs1926447 (p.Ile347Thr), and (**I**) rs3818477 (A>C). (pro)CPB2, (pro)carboxypeptidase B2; C3a, complement 3a; C5a, complement 5a; fbrn, fibrin; flysis, fibrinolysis;

While CPB2 circulates in plasma at a concentration of around approximately 75 nM, it is characterized by a wide reference range in healthy individuals (50–250 nM).[21,22] Although no individuals with a complete CPB2 deficiency have been identified to date, genetic polymorphisms are thought to explain around 25% of the variation in plasma CPB2 levels.[23] CPB2 gene (*CPB2*) polymorphisms have not been investigated in the transplant context; however, since CPB2 has anti-inflammatory as well as anti-fibrinolytic properties, altered circulating CPB2 levels could hypothetically protect or predispose the graft to injury depending on the context.[24–26] Furthermore, considering the contribution of complement activation to allograft injury and dysfunction in kidney transplantation, CPB2 is of further interest because of its modulatory effect on this system.[6,27]

Here, we employed a large cohort of 1,271 renal transplant donors and recipients and leveraged human genetics to determine the role of CPB2 in kidney transplantation. Looking at functional *CPB2* polymorphisms in donor-recipient pairs, and also evaluating potential effect modification by other relevant complement system polymorphisms, we investigated the association between these polymorphisms and long-term graft survival to assess the role of CPB2 on graft survival in kidney transplantation.

## Methods

### Study design and subjects

For this longitudinal cohort study, transplant recipients were enrolled who received a single kidney allograft between March 1993 until February 2008 at the University Medical Center Groningen, as described previously.[28–31] Exclusion criteria were: Lack of DNA, re-transplantation, technical complications during surgery, and loss of follow-up. In brief, a total of 1,271 out of 1,430 donor-recipient kidney transplant pairs were included. All subjects provided written informed consent. The Institutional Review Board approved the study protocol (METc 2014/077), which adhered to the Declaration of Helsinki. The endpoint of our study was graft loss during follow-up with a maximum of 15 years. Graft loss was defined as the need for dialysis or re-transplantation.

### DNA extraction and genotyping

Peripheral blood mononuclear cells were isolated from blood or splenocytes that were collected from both the donor and recipient. DNA was extracted with a commercial kit as instructed by the manufacturer and stored at -80°C. Genotyping of the SNPs was determined via the Illumina VeraCode GoldenGate Assay kit (Illumina, San Diego, CA, USA), according to the manufacturer’s instructions. The promoter of *CPB2* contains several polymorphisms, of which the rs2146881 G > A *CPB2* SNP is the most extensively studied. The minor allele of this polymorphism has been associated with lower plasma levels of CPB2.[32–35] In addition, we chose the rs3742264 G > A (505 G/A, Ala147Thr) and the rs1926447 C > T (1040 C/T, Thr325Ile) *CPB2* SNPs. The minor allele of the rs3742264 is associated with higher plasma levels, whereas the minor allele of the rs1926447 is linked to lower CPB2 levels.[32–35] Lastly, we included the rs3818477 A > C (i4 + 164) *CPB2* SNP, of which the minor allele has previously been shown to associate with lower CPB2 levels as well.[32] Additionally, we genotyped two common functional polymorphism in the C3 and C5 gene: (i) The rs2230199 C>G (Gly102Arg) *C3* SNP and (ii) The rs17611 G>A (2404 G/A, Val802Ile) *C5* SNP.[36,37] We assessed the combinations of the A-allele of the *CPB2* rs3742264 (referred to as the *CPB2*_high_ variant) with the GG-genotype of the *C3* SNP (referred to as C3_102G_ variant) or the AA-genotype of the *C5* SNP (referred to as C5_V802_ variant). Genotype clustering and calling were performed using BeadStudio Software (Illumina).

### Genetic risk score

We compiled a polygenic CPB2 risk score for graft loss based on the presence of the four *CPB2* polymorphisms in the donor and the recipient. To account for the strength of the association of the *CPB2* polymorphisms with graft loss, the score for the presence of a single *CPB2* variant was multiplied by the regression coefficient (that is the logarithm of the hazard ratio) creating a weighted risk score.[38] A regression coefficient is negative when a *CPB2* variant decreases the risk of graft loss and positive when a *CPB2* variant increases the risk of graft loss. The total sum of the protective and hazardous *CPB2* polymorphisms in the donor and recipient combined constitute the value of the CPB2 genetic risk score.

### Statistical analysis

The data are displayed as mean ± standard deviation, median [interquartile ranger (IQR)], or as the total number of patients with percentage [n (%)]. For a two-group comparison, the Mann-Whitney U test, the Student t-test, or the χ^2^ test was used. *CPB2* genotypes were tested for associations with 15-year death-censored graft survival by Kaplan-Meier analysis with log-rank testing. Associations of *CPB2* genotypes with graft loss were further examined by Cox proportional hazards regression analysis with stepwise adjustments for other relevant clinical variables. The models for graft loss included adjustment for recipient demographics (model 2), donor demographics (model 3) or transplant characteristics (model 4). Furthermore, Cox regression models were built with a stepwise forward selection including all variables that significantly associated with graft loss in univariable analysis. Lastly, associations of *CPB2* genotypes with graft loss were verified in subgroups by Cox proportional hazards regression analysis. For continuous variables the subgroups were based on below versus above the median or mean.

We determined Harrell’s concordance statistic (C statistic) to test how well a model distinguished between transplant recipients who developed graft loss and those who did not, additionally taking follow-up into account.[39] As our outcome variable graft loss is dichotomous, the Harrell’s C statistic corresponds to the area under the ROC curve.[40] A value of “1” indicates perfect discrimination, while a value of “0.5” indicates a performance equal to chance. The added value of the *CPB2* genetic risk score to predictive models for graft loss was tested by the integrated discrimination improvement (IDI). The IDI indicates the ability of the new model to improve average sensitivity without decreasing average specificity.[39,41] Statical testing was two-tailed and *P*<0.05 was regarded as significant. Statistical analyses were performed using SPSS software version 25 (SPSS Inc, Chicago, IL, USA) and STATA Statistical Software: Release 17 (StataCorp., College Station, TX, USA).

## Results

### Patient population

The baseline demographics of the 1,271 donor-recipient pairs, including transplant characteristics are provided in Table 1. During a mean follow-up period of 6.2 ± 4.2 years, 215 kidney transplant recipients lost their grafts (16.9%), whereas 191 recipients died with a functioning graft (15.0%). Deceased donor kidney transplantation, absence of cyclosporin and corticosteroids for immunosuppression, as well as donor and recipient blood type AB were significantly more prevalent in patients who progressed to graft loss. Furthermore, on average patients who developed graft loss were younger, while their donors were typically older, and both cold as well as warm ischemia times were significantly longer. Lastly, delayed graft function occurred more frequently in patients who experienced graft loss. The above-mentioned clinical characteristics were all significantly associated with graft loss in univariable analyses (Table 1).

**Table 1:**
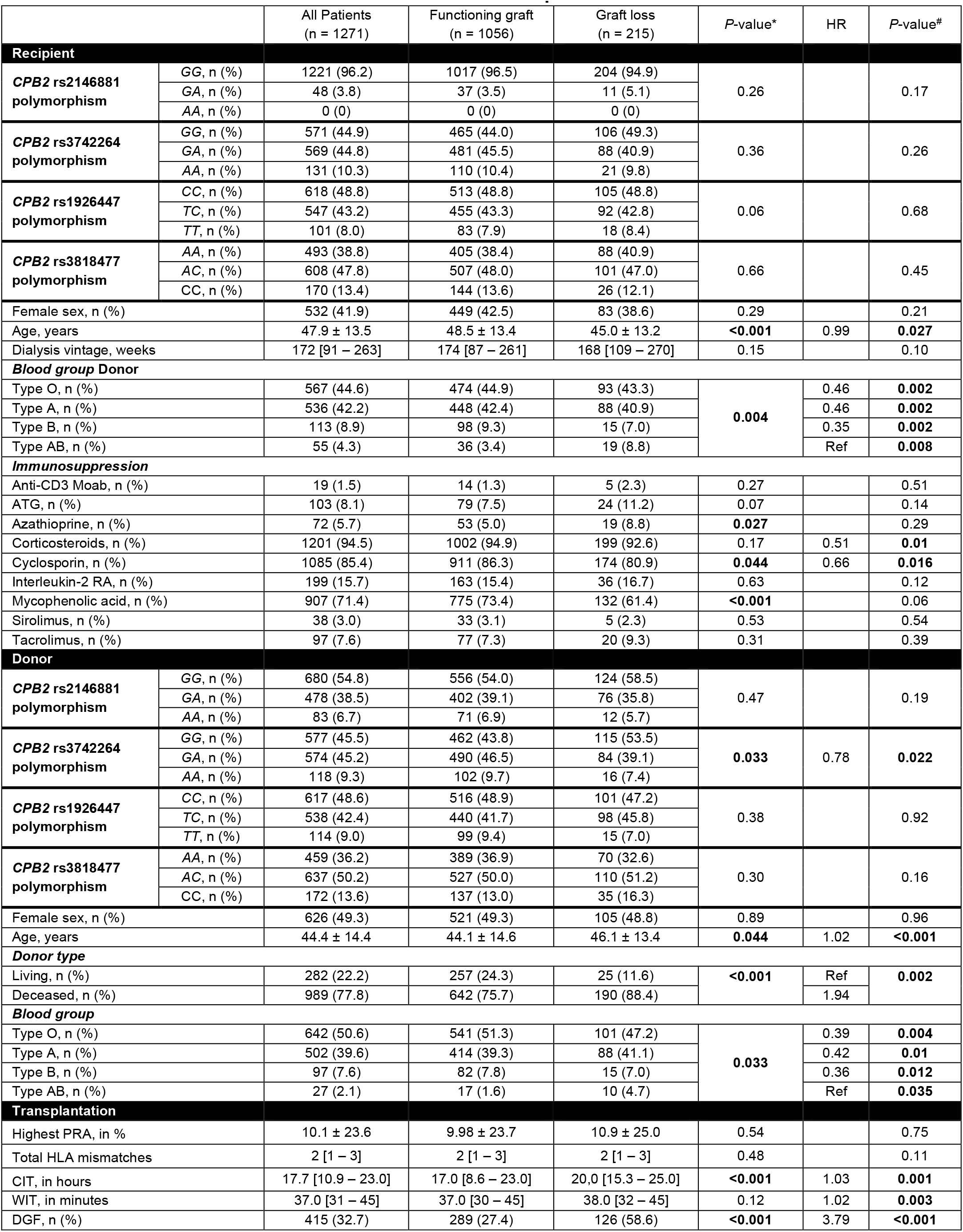
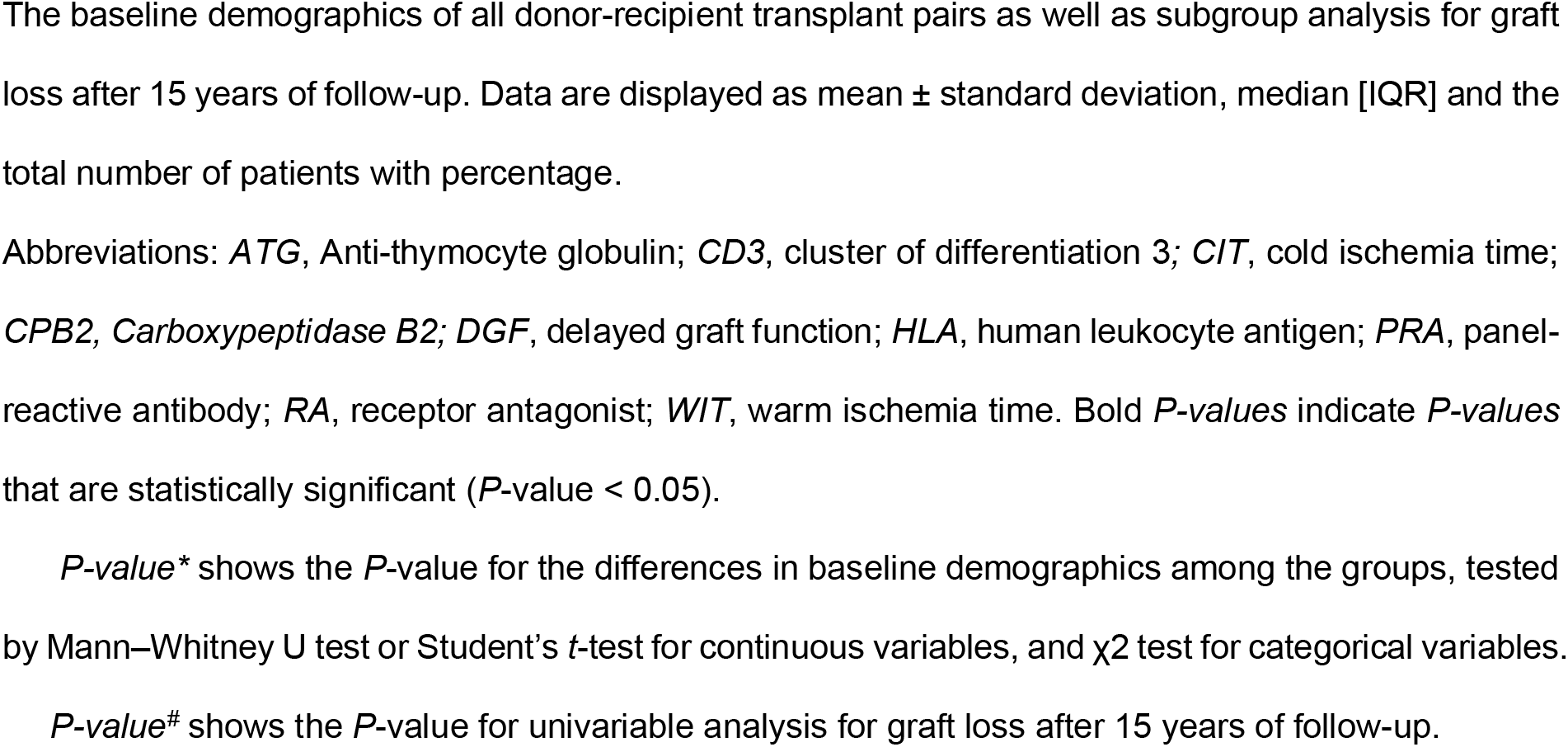
Baseline characteristics of the donors and recipients.

|  |  | All Patients<br>(n = 1271) | Functioning graft<br>(n = 1056) | Graft loss<br>(n = 215) | P-value* | HR | P-value# |
| --- | --- | --- | --- | --- | --- | --- | --- |
| Recipient |  |  |  |  |  |  |  |
| CPB2 rs2146881 polymorphism | GG, n (%) | 1221 (96.2) | 1017 (96.5) | 204 (94.9) | 0.26 |  | 0.17 |
|  | GA, n (%) | 48 (3.8) | 37 (3.5) | 11 (5.1) |  |  |  |
|  | AA, n (%) | 0 (0) | 0 (0) | 0 (0) |  |  |  |
| CPB2 rs3742264 polymorphism | GG, n (%) | 571 (44.9) | 465 (44.0) | 106 (49.3) | 0.36 |  | 0.26 |
|  | GA, n (%) | 569 (44.8) | 481 (45.5) | 88 (40.9) |  |  |  |
|  | AA, n (%) | 131 (10.3) | 110 (10.4) | 21 (9.8) |  |  |  |
| CPB2 rs1926447 polymorphism | CC, n (%) | 618 (48.8) | 513 (48.8) | 105 (48.8) | 0.06 |  | 0.68 |
|  | TC, n (%) | 547 (43.2) | 455 (43.3) | 92 (42.8) |  |  |  |
|  | TT, n (%) | 101 (8.0) | 83 (7.9) | 18 (8.4) |  |  |  |
| CPB2 rs3818477 polymorphism | AA, n (%) | 493 (38.8) | 405 (38.4) | 88 (40.9) | 0.66 |  | 0.45 |
|  | AC, n (%) | 608 (47.8) | 507 (48.0) | 101 (47.0) |  |  |  |
|  | CC, n (%) | 170 (13.4) | 144 (13.6) | 26 (12.1) |  |  |  |
| Female sex, n (%) |  | 532 (41.9) | 449 (42.5) | 83 (38.6) | 0.29 |  | 0.21 |
| Age, years |  | 47.9 ± 13.5 | 48.5 ± 13.4 | 45.0 ± 13.2 | <0.001 | 0.99 | 0.027 |
| Dialysis vintage, weeks |  | 172 [91 – 263] | 174 [87 – 261] | 168 [109 – 270] | 0.15 |  | 0.10 |
| Blood group Donor |  |  |  |  |  |  |  |
| Type O, n (%) |  | 567 (44.6) | 474 (44.9) | 93 (43.3) | 0.004 | 0.46 | 0.002 |
| Type A, n (%) |  | 536 (42.2) | 448 (42.4) | 88 (40.9) |  | 0.46 | 0.002 |
| Type B, n (%) |  | 113 (8.9) | 98 (9.3) | 15 (7.0) |  | 0.35 | 0.002 |
| Type AB, n (%) |  | 55 (4.3) | 36 (3.4) | 19 (8.8) |  | Ref | 0.008 |
| Immunosuppression |  |  |  |  |  |  |  |
| Anti-CD3 Moab, n (%) |  | 19 (1.5) | 14 (1.3) | 5 (2.3) | 0.27 |  | 0.51 |
| ATG, n (%) |  | 103 (8.1) | 79 (7.5) | 24 (11.2) | 0.07 |  | 0.14 |
| Azathioprine, n (%) |  | 72 (5.7) | 53 (5.0) | 19 (8.8) | 0.027 |  | 0.29 |
| Corticosteroids, n (%) |  | 1201 (94.5) | 1002 (94.9) | 199 (92.6) | 0.17 | 0.51 | 0.01 |
| Cyclosporin, n (%) |  | 1085 (85.4) | 911 (86.3) | 174 (80.9) | 0.044 | 0.66 | 0.016 |
| Interleukin-2 RA, n (%) |  | 199 (15.7) | 163 (15.4) | 36 (16.7) | 0.63 |  | 0.12 |
| Mycophenolic acid, n (%) |  | 907 (71.4) | 775 (73.4) | 132 (61.4) | <0.001 |  | 0.06 |
| Sirolimus, n (%) |  | 38 (3.0) | 33 (3.1) | 5 (2.3) | 0.53 |  | 0.54 |
| Tacrolimus, n (%) |  | 97 (7.6) | 77 (7.3) | 20 (9.3) | 0.31 |  | 0.39 |
| Donor |  |  |  |  |  |  |  |
| CPB2 rs2146881 polymorphism | GG, n (%) | 680 (54.8) | 556 (54.0) | 124 (58.5) | 0.47 |  | 0.19 |
|  | GA, n (%) | 478 (38.5) | 402 (39.1) | 76 (35.8) |  |  |  |
|  | AA, n (%) | 83 (6.7) | 71 (6.9) | 12 (5.7) |  |  |  |
| CPB2 rs3742264 polymorphism | GG, n (%) | 577 (45.5) | 462 (43.8) | 115 (53.5) | 0.033 | 0.78 | 0.022 |
|  | GA, n (%) | 574 (45.2) | 490 (46.5) | 84 (39.1) |  |  |  |
|  | AA, n (%) | 118 (9.3) | 102 (9.7) | 16 (7.4) |  |  |  |
| CPB2 rs1926447 polymorphism | CC, n (%) | 617 (48.6) | 516 (48.9) | 101 (47.2) | 0.38 |  | 0.92 |
|  | TC, n (%) | 538 (42.4) | 440 (41.7) | 98 (45.8) |  |  |  |
|  | TT, n (%) | 114 (9.0) | 99 (9.4) | 15 (7.0) |  |  |  |
| CPB2 rs3818477 polymorphism | AA, n (%) | 459 (36.2) | 389 (36.9) | 70 (32.6) | 0.30 |  | 0.16 |
|  | AC, n (%) | 637 (50.2) | 527 (50.0) | 110 (51.2) |  |  |  |
|  | CC, n (%) | 172 (13.6) | 137 (13.0) | 35 (16.3) |  |  |  |
| Female sex, n (%) |  | 626 (49.3) | 521 (49.3) | 105 (48.8) | 0.89 |  | 0.96 |
| Age, years |  | 44.4 ± 14.4 | 44.1 ± 14.6 | 46.1 ± 13.4 | 0.044 | 1.02 | <0.001 |
| Donor type |  |  |  |  |  |  |  |
| Living, n (%) |  | 282 (22.2) | 257 (24.3) | 25 (11.6) | <0.001 | Ref | 0.002 |
| Deceased, n (%) |  | 989 (77.8) | 642 (75.7) | 190 (88.4) |  | 1.94 |  |
| Blood group |  |  |  |  |  |  |  |
| Type O, n (%) |  | 642 (50.6) | 541 (51.3) | 101 (47.2) | 0.033 | 0.39 | 0.004 |
| Type A, n (%) |  | 502 (39.6) | 414 (39.3) | 88 (41.1) |  | 0.42 | 0.01 |
| Type B, n (%) |  | 97 (7.6) | 82 (7.8) | 15 (7.0) |  | 0.36 | 0.012 |
| Type AB, n (%) |  | 27 (2.1) | 17 (1.6) | 10 (4.7) |  | Ref | 0.035 |
| Transplantation |  |  |  |  |  |  |  |
| Highest PRA, in % |  | 10.1 ± 23.6 | 9.98 ± 23.7 | 10.9 ± 25.0 | 0.54 |  | 0.75 |
| Total HLA mismatches |  | 2 [1 – 3] | 2 [1 – 3] | 2 [1 – 3] | 0.48 |  | 0.11 |
| CIT, in hours |  | 17.7 [10.9 – 23.0] | 17.0 [8.6 – 23.0] | 20.0 [15.3 – 25.0] | <0.001 | 1.03 | 0.001 |
| WIT, in minutes |  | 37.0 [31 – 45] | 37.0 [30 – 45] | 38.0 [32 – 45] | 0.12 | 1.02 | 0.003 |
| DGF, n (%) |  | 415 (32.7) | 289 (27.4) | 126 (58.6) | <0.001 | 3.79 | <0.001 |

The baseline demographics of all donor-recipient transplant pairs as well as subgroup analysis for graft loss after 15 years of follow-up. Data are displayed as mean ± standard deviation, median [IQR] and the total number of patients with percentage.
Abbreviations: *ATG*, Anti-thymocyte globulin; *CD3*, cluster of differentiation 3; *CIT*, cold ischemia time; *CPB2*, Carboxypeptidase B2; *DGF*, delayed graft function; *HLA*, human leukocyte antigen; *PRA*, panel-reactive antibody; *RA*, receptor antagonist; *WIT*, warm ischemia time. Bold *P-values* indicate *P-values* that are statistically significant ( $P$ -value < 0.05).
*P-value*<sup>\*</sup> shows the *P*-value for the differences in baseline demographics among the groups, tested by Mann–Whitney U test or Student's *t*-test for continuous variables, and $\chi^2$ test for categorical variables.
*P-value*<sup>#</sup> shows the *P*-value for univariable analysis for graft loss after 15 years of follow-up.

We genotyped four common polymorphisms of the *CPB2* (Fig. 1F – I): (i) the rs2146881 G > A in the promoter region at position ∼438 (Fig. 1F), (ii) the rs3742264 G > A encoding for a Thr to Ala substitution at position 147 (Fig. 1G), (iii) the rs1926447 C > T resulting in a Thr to Ile substitution at position 325 (Fig. 1H), and (iv) rs3818477 A > C in intron 4 (Fig. 1I). The observed genotypic frequencies are shown in Table 1. For the *CPB2* rs2146881 G > A SNP, the frequency of the minor allele was significantly lower in recipients compared to donors (*P* < 0.0001) and to the European cohort of the 1000 genomes project (*P* < 0.0001, supplementary data). Furthermore, the frequencies of the *CPB2* rs3818477 A > C SNP in the donors significantly differed from those reported by the European cohort of the 1,000 genomes project (*P* = 0.02), but not compared to recipients (*P* = 0.39, supplementary data). No differences were seen in genotypic frequencies between the donors, recipients, and the European cohort of 1,000 genomes project for the other CPB2 polymorphisms (supplementary data). The distribution of all four polymorphisms was in Hardy−Weinberg equilibrium.

### A high-producing *CPB2* variant associates with graft loss after kidney transplantation

We first examined whether polymorphisms in *CPB2* associated with graft loss after kidney transplantation. Of all the assessed *CPB2* genetic variants, only the rs3742264 polymorphism in the donor significantly associated with 15-year death-censored graft survival (*P* = 0.022). Kaplan–Meier survival analysis revealed that the minor allele of the *CPB2* rs3742264 SNP in the donor was associated with a lower risk of graft loss after transplantation (Fig. 2A, *P* = 0.044). The cumulative incidence of graft loss after 15 years of follow-up was 19.9% in the reference GG-genotype group, 14.6% in the GA-genotype group, and 13.3% in the AA-genotype group, respectively. For further analysis the GA- and AA-genotype were combined to one group (Fig. 2B, *P* = 0.013), since the incidence of graft loss was not significantly different between these genotypes. In univariable analysis, the minor allele of the *CPB2* rs3742264 SNP in the donor was significantly associated with improved 15-year death-censored graft survival (HR, 0.71; 95%-CI, 0.55 – 0.93; *P* = 0.014). Multivariable models were constructed using a stepwise forward selection procedure including all variables that were significantly associated with graft loss in the univariable analysis (Table 2). In the final model, the *CPB2* rs3742264 SNP in the donor, recipient age, the occurrence of delayed graft function, recipient blood type, and donor age were included. After adjustment, the minor allele of the *CPB2* rs3742264 SNP in the donor was significantly associated with a reduced risk of graft loss (HR, 0.67; 95% CI: 0.51 – 0.88, *P* = 0.004).

**Figure 2:**
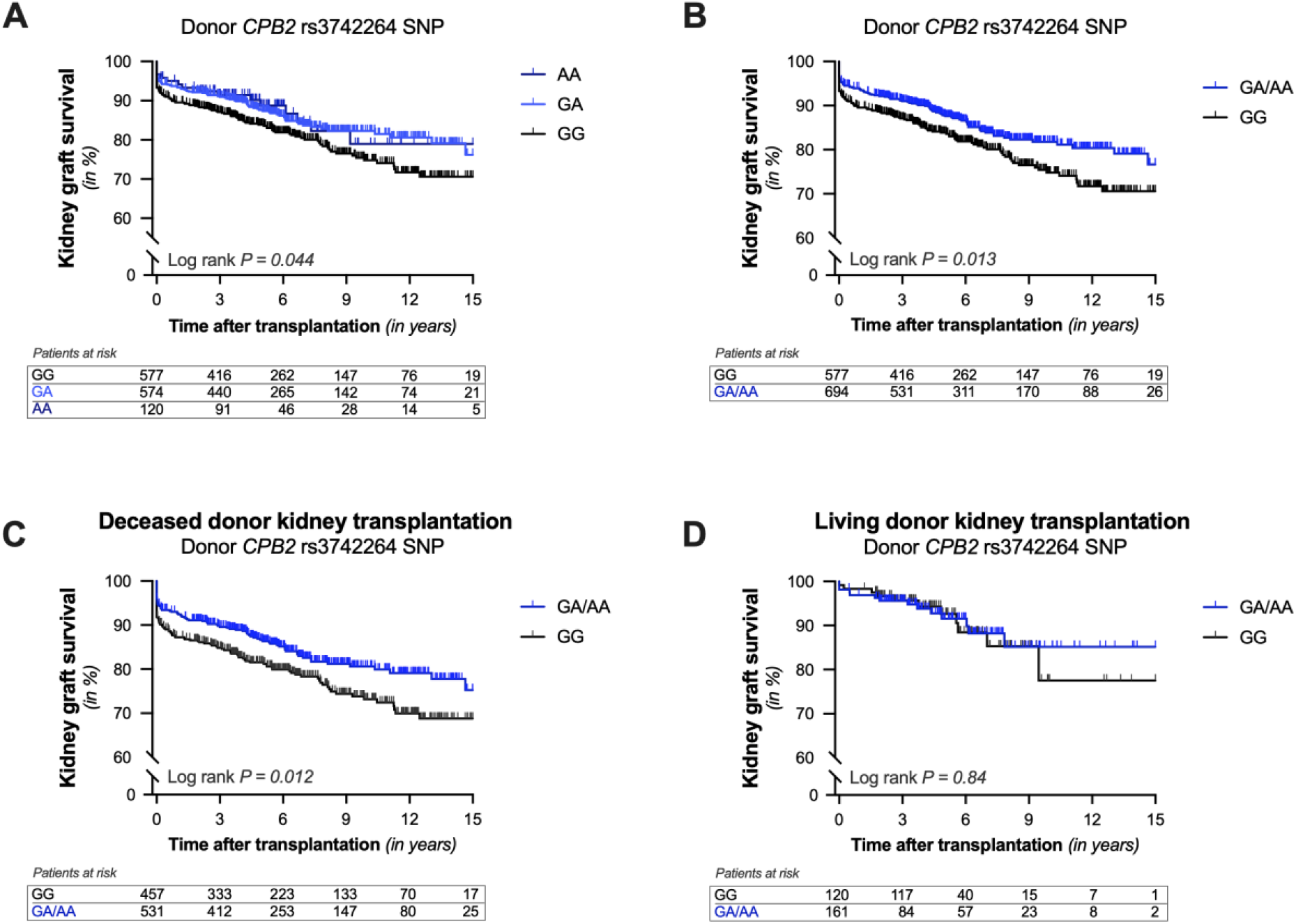
Kaplan-Meier curves for 15-year death-censored kidney graft survival according to the presence of a carboxypeptidase B2 gene polymorphism in the donor. **(A)** Cumulative 15-year death-censored kidney graft survival with the rs3742264 G>A polymorphism in the carboxypeptidase B2 gene (*CBP2*) in the donor. (**B**) Next, the GA- and AA-genotype were combined in one group since the graft loss incidence did not significantly differ between these genotypes. A subgroup analysis for donor type was performed. Cumulative 15-year death-censored kidney graft survival with the rs3742264 G>A polymorphism in (**C**) deceased kidney donors and (**D**) living kidney donors. The Log-rank test was used to compare the graft loss incidence between the different groups.

**Table 2:** Multivariable analysis of 15-year death-censored graft survival.

| Variables not in the equation |  | Variables in the equation |  |  |
| --- | --- | --- | --- | --- |
| *Variables* | *P-value* | *Variables* | *P-value* | *Hazard Ratio* |
| Warm ischemia time<br>(minutes) | 0.07 | rs3742264 in the donor<br>(G versus A) | 0.004 | 0.67 (1.21 – 3.82) |
| Corticosteroids | 0.09 | Recipient age<br>(years) | <0.001 | 0.98 (0.97 – 0.99) |
| Cold ischemia time<br>(hours) | 0.11 | Delayed graft function<br>(yes versus no) | <0.001 | 4.02 (3.03 – 5.32) |
| Donor type<br>(living versus deceased) | 0.13 | Recipient blood type<br>(ABO versus other) | 0.001 |  |
| Cyclosporin | 0.35 | Donor age<br>(years) | 0.002 | 1.02 (1.01 – 1.03) |
| Donor blood type<br>(ABO versus other) | 0.98 |  |  |  |

Multivariable cox regression was performed with a stepwise forward selection. Only variables that with a *P*-value < 0.05 in the univariate analysis were included. Data are presented as hazard ratio with 95% confidence interval (CI) and *P*-value. In the final model, the *CPB2* SNP (rs3742264-T) in the donor, recipient age, the occurrence of delayed graft function, recipient blood type, and donor age were included, whereas warm ischemia time, use of corticosteroids, cold ischemia time, donor type, use of cyclosporin, and donor blood type were not.

Furthermore, we performed a subgroup analysis for the donor type, since deceased organ donors are often characterized by coagulopathies.[42] Kaplan-Meier curves demonstrated that the association remained significant between the *CPB2* rs3742264 SNP in the donor and long-term graft survival for kidney allografts from deceased donors (Fig. 2C, *P* = 0.012). The association, however, lost statistical significance for kidney allografts from living donors (Fig. 2D, *P* = 0.84). Taken together, our results show that the high-producing *CPB2* rs3742264 polymorphism in deceased organ donors is associated with a reduced risk of graft loss after kidney transplantation.

### *CPB2* associates with graft loss potentially through inactivation of complement anaphylatoxins

Next, we investigated if the association between the high-producing *CPB2* rs3742264 SNP and long-term graft survival was mediated via CPB2 ability to inactivate complement anaphylatoxin C3a and C5a. On this basis, we tested the combined effect of the high-producing *CPB2* variant with gain-of-function complement polymorphisms on graft loss after kidney transplantation.

First, we combined the high-producing *CPB2* polymorphism with a common functional polymorphism in C3 gene (*C3*) rs2230199 resulting in a glycine to arginine substitution at position 102. The minor allele of this polymorphism results in a *C3* variant (C3_102G_) that is less well inhibited, leading to increased C3 activation and higher C3a formation.[43] Next, donor-recipient pairs were separated into four groups according to the presence or absence of the *C3* and *CPB2* polymorphism in the donor. Kaplan–Meier survival analyses showed a significant difference in graft failure rates among the four groups (*P* = 0.022, Fig. 3A). Kidney allografts possessing the *CPB2*_high_ variant and the reference *C3*_102R_ variant had the best outcome (15-year death-censored graft survival: 77.2%), whereas kidney allografts possessing the *C3*_102G_ variant and the refence *CPB2*_normal_ variant had the worst outcome (15-year death-censored graft survival: 50.2%). Moreover, kidney allografts possessing either both variants or no variants showed similar outcomes, suggesting that the protective effect of the *CPB2*_high_ variant could be mitigated by the hazardous effect of the *C3*_102R_ variant.

**Figure 3:**
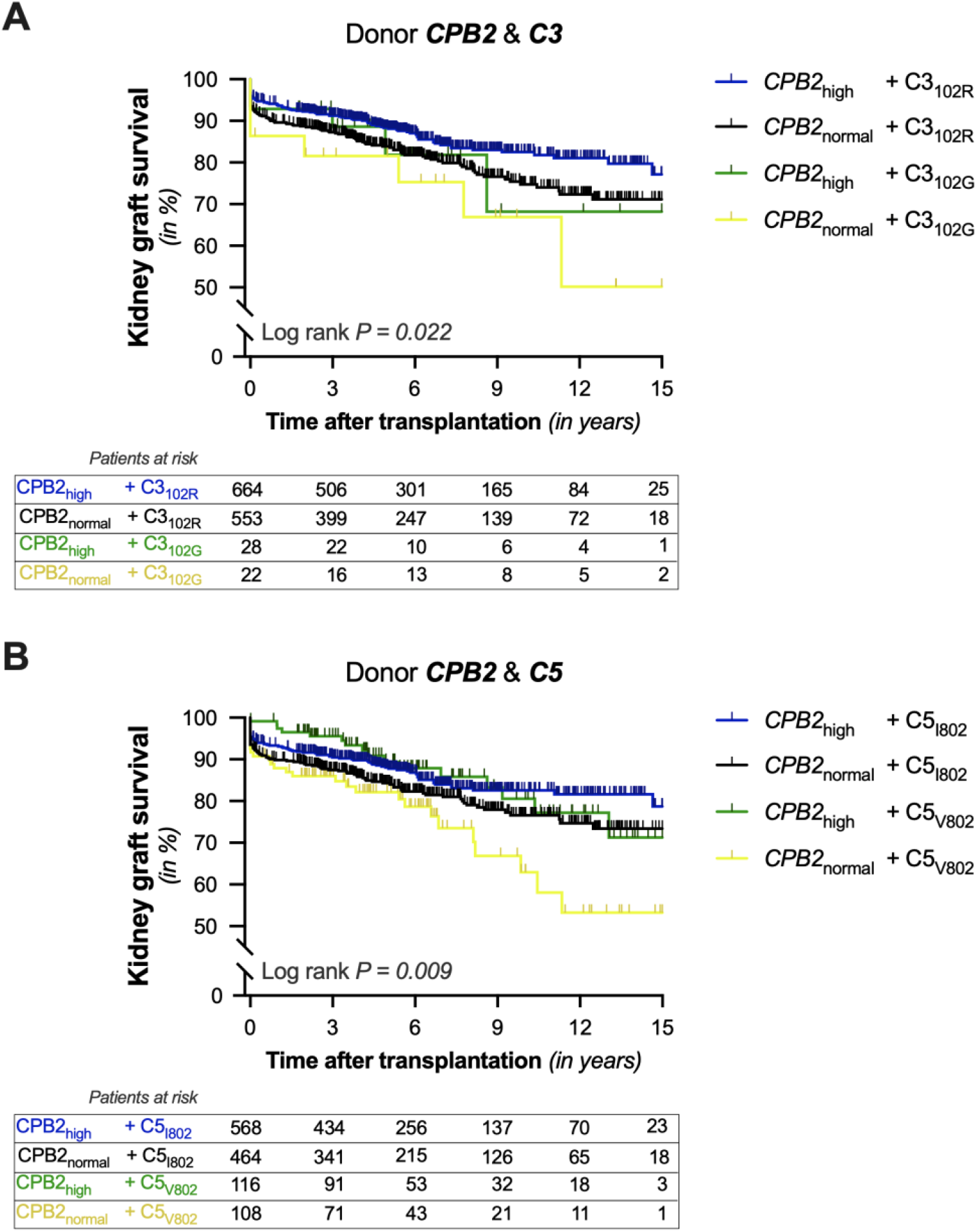
Kaplan-Meier curves for 15-year death-censored kidney graft survival according to the presence of a carboxypeptidase B2, C3 and C5 gene polymorphism in the donor. **(A)** Cumulative 15-year death-censored kidney graft survival according to the presence of the rs3742264 G>A polymorphism in the carboxypeptidase B2 gene (*CBP2*) and the rs2230199 C > G polymorphism in the C3 gene (*C3*). Pairs were divided into four groups according to the absence (black line), presence of the CPB2 variant (blue line), presence of the C3 variant (yellow line) or both (green line). (**B**) Cumulative 15-year death-censored kidney graft survival according to the presence of the rs3742264 G>A polymorphism in the carboxypeptidase B2 gene (*CBP2*) and the rs17611 G > A polymorphism in the C5 gene (*C5*). Pairs were divided into four groups according to the absence (black line), presence of the CPB2 variant (blue line), presence of the C5 variant (yellow line) or both (green line). Log-rank test was used to compare the incidence of graft loss between the groups.

**Figure 4:**
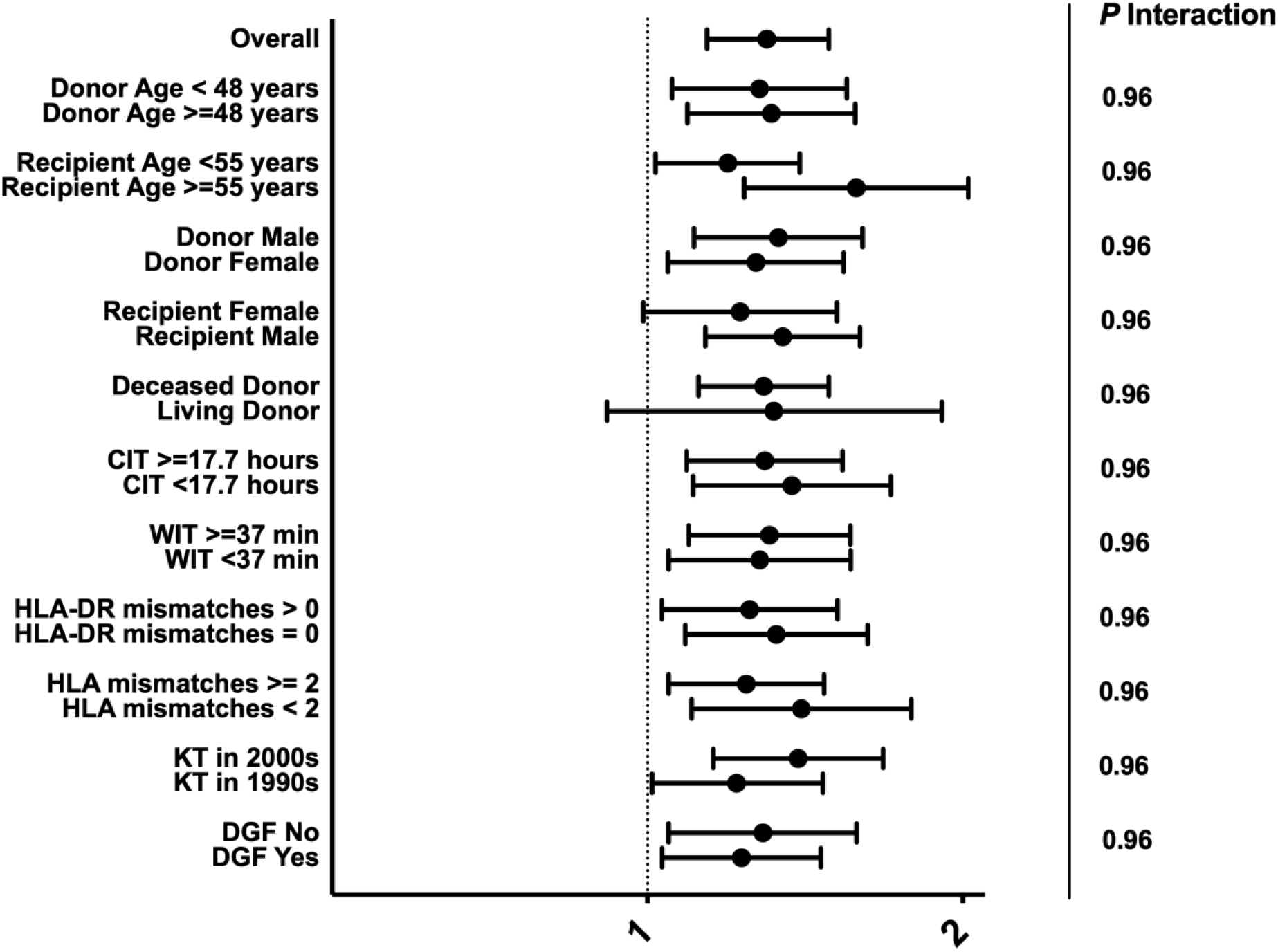
**Hazard ratios for CPB2 genetic risk score among subgroups.** Forest plot of CPB2 genetic risk score sub-analyses, demonstrating consistency of the hazard ratios for graft loss in the different subgroups with the exception of donor origins of the kidney allografts and recipient sex. The association between the CPB2 genetic risk score and graft loss was not observed in kidney transplants from living donors or in female recipients. No significant interaction was seen between the CPB2 genetic risk score and the various clinical variables of the subgroups.

Next, we combined the high-producing *CPB2* polymorphism with a common functional polymorphism in C5 gene (*C5*) rs17611 resulting in a valine to isoleucine substitution at position 802. The G-allele of this polymorphism results in a *C5* variant (C5_V802_) that is more susceptible to cleavage, leading to enhanced C5a production.[44] Once again, donor-recipient pairs were separated into four groups according to the presence or absence of the *C5* and *CPB2* polymorphism in the donor. Kaplan– Meier survival analyses showed a significant difference in graft failure rates among the four groups (*P* = 0.009, Fig. 3B). Kidney allografts possessing the *CPB2*_high_ variant and the reference *C5*_I802_ variant had the best outcome (15-year death-censored graft survival: 78.7%), whereas kidney allografts possessing the *C5*_V802_ variant and the refence *CPB2*_normal_ variant had the worst outcome (15-year death-censored graft survival: 53.2%). Moreover, kidney allografts possessing either both variants or no variants experienced similar outcomes, suggesting once again the protective effect of the *CPB2*_high_ variant could be mitigated by the hazardous effect of the *C5*_V802_ variant. In conclusion, our data implies that the association of the high-producing *CPB2* rs3742264 SNP with graft loss is possibly mediated via CPB2’s ability to inactivate complement anaphylatoxin C3a and C5a.

### A CPB2 genetic risk score improves risk prediction for graft loss

In addition to analyzing the *CPB2* polymorphisms separately, we also compiled a genetic risk score of the four *CPB2* variants in both the recipients and donors. Each polymorphism was weighted based on its hazard ratio, creating a positive coefficient for hazardous SNPs and a negative one for protective SNPs. Overall, a genetic risk score below zero indicates the presence of more protective *CPB2* SNPs in the donor-recipient renal transplant pairs, while a genetic risk score above zero alludes to more hazardous *CPB2* SNPs in a transplant pair.

The CPB2 genetic risk score significantly associated with 15-year death-censored graft survival in univariable analysis (HR, 1.30; 95%-CI, 1.14 – 1.49; *P*<0.001 per SD increase). Multivariable analysis was performed with stepwise adjustments for additional relevant clinical variables (Table 3), including recipient characteristics (model 2), donor characteristics (model 3), and transplant variables (model 4). In Cox regression analysis, the CPB2 genetic risk score remained significantly associated with graft loss independent of potential confounders. Thereafter, we investigated whether the CPB2 genetic risk score (including the *CPB2* rs3742264 SNP) was a better predictor of graft loss than the *CPB2* rs3742264 SNP in the donor alone by multivariable regression with a stepwise forward selection (supplementary data). In the final model, the CPB2 genetic risk score containing the *CPB2* rs3742264 SNP was included, whereas the single *CPB2* rs3742264 SNP in the donor was excluded. After adjustment, the CPB2 genetic risk score was associated with graft loss with a hazard ratio of 1.31 per SD increase (95%-CI: 1.14 – 1.50; *P*<0.001). In addition, the hazard ratio of the CPB2 genetic risk score was consistent and remained significant in subgroup analyses (Figure 3); except for living donors and in female recipients. The 95%-CI of the various subgroups had substantially overlapped with the general hazard ratio, signifying the consistency of the impact of the CPB2 genetic risk score across the subgroups.

**Table 3:** Associations of CPB2 genetic risk score with graft loss.

| CPB2 genetic risk score |  |  |  |
| --- | --- | --- | --- |
|  | Hazard ratio<br>(per SD) | 95% CI | P-value |
| **Model 1** | 1.302 | 1.140 – 1.488 | <0.001 |
| **Model 2** | 1.309 | 1.139 – 1.505 | <0.001 |
| **Model 3** | 1.279 | 1.118 – 1.463 | <0.001 |
| **Model 4** | 1.268 | 1.102 – 1.458 | 0.001 |

Data are presented as the hazard ratio with a 95% confidence interval (CI) and P-value.
**Model 1:** Crude model.
**Model 2:** Adjusted for model 1 plus recipient characteristic's: recipient age, recipient sex, recipient blood type and dialysis vintage.
**Model 3:** Adjusted for model 1 plus donor characteristic's: donor age, donor sex, donor blood type, and donor origin.
**Model 4:** Adjusted for model 1 plus transplant characteristic's: cold and warm ischemia time, the total HLA-mismatches, and the occurrence of delayed graft function (DGF).

Finally, the performance of the CPB2 genetic risk score for the prediction of graft loss was also assessed (Table 4). The CPB2 genetic risk score had a Harrell’s C of 0.59 (95% CI: 0.55 – 0.63). Moreover, when added to a model of the *CPB2* s3742264 SNP in the donor (c-statistic, 0.55; 95%-CI, 0.51 – 0.59), the CPB2 genetic risk score significantly improved the Harrell’s C (c-statistic increase, 0.035; 95%-CI, 0.005 – 0.066; *P* = 0.02). As additional variables were included and the discriminative accuracy to predict graft loss of the model improved. The Harrell’s C of the models with the recipient, donor and transplant characteristics significantly increased with the addition of the CPB2 genetic risk score (Model 3 – 5), while only a trend was seen in the model with all variables that were significantly associated with graft loss in multivariable regression analysis (Model 6). Lastly, the CPB2 genetic risk score significantly increased the predictive value of the models according to the IDI.

**Table 4:** Additive value of the *CPB2* genetic risk score for the prediction of graft loss

|  | Harrell's C (95% CI) |  | Change (95%-CI)* | P-value | IDI (%) | P-value |
| --- | --- | --- | --- | --- | --- | --- |
|  | Without the CPB2 genetic risk score | with the CPB2 genetic risk score |  |  |  |  |
| **Model 1** | 0.500 | 0.588 (0.546 – 0.629) | N/A | N/A | N/A | N/A |
| **Model 2** | 0.550 (0.514 – 0.586) | 0.585 (0.544 – 0.626) | 0.035 (0.005 – 0.066) | 0.02 | 0.7 | 0.003 |
| **Model 3** | 0.566 (0.525 – 0.607) | 0.615 (0.575 – 0.656) | 0.051 (0.016 – 0.086) | 0.004 | 1.2 | 0.001 |
| **Model 4** | 0.623 (0.587 – 0.660) | 0.645 (0.608 – 0.682) | 0.028 (0.003 – 0.053) | 0.03 | 1.0 | 0.003 |
| **Model 5** | 0.704 (0.668 – 0.740) | 0.722 (0.685 – 0.759) | 0.021 (0.000 – 0.042) | 0.06 | 1.0 | 0.005 |
| **Model 6** | 0.734 (0.700 – 0.769) | 0.744 (0.711 – 0.777) | 0.012 (-0.001 – 0.025) | 0.08 | 1.1 | 0.007 |

*Data are presented as Harrell's concordance statistic (Harrell's C) with a 95% confidence interval (CI)* *and integrated discrimination improvement (IDI) with P-value. \*Change in C-statistics compared to the* *model without the CPB2.*
**Model 1:** Crude model.
**Model 2:** CPB2 rs3742264 polymorphism in the donor
**Model 3:** Adjusted for recipient characteristic's: recipient age, recipient sex, recipient blood type and dialysis vintage.
**Model 4:** Adjusted for donor characteristic's: donor age, donor sex, donor blood type, and donor origin.
**Model 5:** Adjusted for plus transplant characteristic's: cold and warm ischemia time, the total HLA- mismatches, and the occurrence of delayed graft function.
**Model 6:** Adjusted for the occurrence of delayed graft function, recipient blood type, donor age and use of corticosteroids.

## Discussion

Here, we report that kidney transplantation from allografts possessing a high-producing *CPB2* polymorphism results in a significantly attenuated risk of graft failure. We propose that the higher survival rate could be explained by the ability of CPB2 to inactive complement anaphylatoxins (i.e., C3a and C5a), since the protective effect of the *CPB2* polymorphism on graft loss could be reversed by gain-of-function polymorphisms in complement genes (i.e. *C3* and *C5*). Furthermore, we demonstrated that a genetic risk score based on four CPB2 polymorphisms in the donor as well as the recipient was a important determinant of long-term graft survival. The present data also proposes CPB2 as a novel potential therapeutic strategy to improve kidney transplantation outcomes, especially since therapeutic strategies informed by human genetic evidence are more than twice as likely to lead to approved therapeutics.[45,46]

CPB2, because of its anti-fibrinolytic activity, has predominantly been studied in cardiovascular disease and thrombotic disorders.[22] Increased levels of CPB2 are anticipated to induce a hypofibrinolytic state and thereby represent a potential risk factor for various coagulation disorders. Accordingly, thrombus formation was shown to be ameliorated in mice that are deficient in CPB2 (*Cpb2*^-/-^) upon ferric chloride (FeCl_3_)-induced vena cava thrombosis.[47] Numerous studies have subsequently used human genetics to investigate the role of CPB2 in thromboembolic diseases by evaluating the association between *CPB2* polymorphisms and disease risk.[22] Recently, a meta-analysis of 23 studies investigated the impact of *CPB2* variants on the risk of cardiovascular disease, failing to confirm a significant association.[48] Nevertheless, in sub-analyses an increased risk of cardiovascular disease was detected with a certain *CPB2* polymorphisms, suggesting an intricate relationship between CPB2 and thrombotic disease.

Apart from having anti-fibrinolytic activity during clot formation, CPB2 also has an anti-inflammatory role.[10] These anti-inflammatory properties are mediated through the cleavage of C-terminal arginine from pro-inflammatory mediators such as complement anaphylatoxins, bradykinin, osteopontin, and vascular endothelial growth factor-A.[10,17–20] Recent *In-vivo* evidence by Song *and collaegues,* demonstrates that CPB2 dampened inflammation in autoimmune arthritis.[49] After the induction of anti-collagen antibody-induced arthritis, *Cpb2*^−/−^ mice exhibited more severe arthritis, whereas *C5*^−/−^ mice were protected against arthritis.[49] Intriguingly, an anti-C5 antibody prevented the development of severe arthritis in *Cpb2*^−/−^ mice. In line with these results, *Cpb2*^−/−^ mice also had a greater complement-mediated influx of inflammatory cells in a model of zymosan-induced peritonitis [49]. Furthermore, rheumatoid arthritis patients carrying a *CPB2* variant, resulting in a longer half-life of CPB2, were also less likely to develop severe disease.[49] Altogether, these studies reveal the profound ability of CPB2 to reduce inflammation, especially through the inactivation of complement anaphylatoxins.

To our knowledge, our study is the first to explore the role of CPB2 in kidney transplantation in relation to graft survival. Plasma levels of CPB2 have previously been reported to be elevated in kidney transplant recipients compared to healthy controls.[50,51] Consequently, CPB2 has been postulated to contribute to the hypofibrinolysis seen in this patient population, thereby negatively impacting transplant outcome. Micro-thrombi are frequently formed within the graft during kidney transplantation, which can then lead to poor initial kidney graft function.[42,52,53] Despite these conjectures, however, we found that the rs3742264 *CPB2* polymorphism in kidney allografts reduced the risk of graft failure. Particularly, this *CPB2* variant has previously been associated with higher levels.[32–35] Hence, we hypothesized that the association found was mediated by CPB2 ability to inactivate C3a and C5a, especially since complement anaphylatoxins are known to impact graft outcome in kidney transplantation.[6,36,54–57] To test this, we looked at the combined impact of the *CPB2* variant with *C3* or *C5* polymorphisms that have been reported to lead to higher C3a and C5a levels.[43,44] In lin with our hypothesis, we found kidney allografts possessing the CPB2 variant as well as a complement gain-of-function had almost identical outcomes compared to kidney allografts carrying neither variant. We, therefore, propose that in kidney transplantation, high CPB2 levels improve graft survival by reducing complement-mediated inflammation.

CPN and CPB2 are the only two carboxypeptidases present in plasma.[10] The current paradigm is that carboxypeptidase N (CPN) is the main regulator of C3a and C5a, whereas CPB2 would be primarily involved in fibrinolysis. However, *in-vitro* experiments revealed the ability of CPB2 to inactivate C3a and C5a.[19] Moreover, CPN is less efficient in cleaving C5a into C5a_DesArg_ than CPB2, whereas for C3a both enzymes were equally efficient.[19] Moreover, studies using *Cpb2*^−/−^ mice confirmed that inactivation of complement anaphylatoxins is regulated by CPB2 *in-vivo*. In a model of hemolytic uremic syndrome (HUS), the disease was exacerbated in *Cpb2*^−/−^ mice, compared to both controls and *Cpn*^−/−^ mice.[17] In addition, *Cpb2*^−/−^ mice presented with the clinical HUS triad of thrombocytopenia, uremia, and microangiopathic hemolytic anemia.[17] The exacerbated disease in *Cpb2*^−/−^ mice was attenuated by the treatment with an anti-C5 antibody, and survival after treatment was comparable to wild-type mice.[17] Altogether, our study adds to a growing body of evidence that shows that implicates CPB2 as a chief regulator of C3a and C5a.

Regarding donor-recipient nuances, we found an association between a *CPB2* polymorphism in donors with the risk of graft loss, but not recipients. These results reveal that it is not CPB2 production by the recipient, but instead the local CPB2 production by the donor kidney that positively impacts graft survival in kidney transplantation. Furthermore, when we performed a subgroup analysis for donor type, we observed a significant association between the CPB2 polymorphism in deceased donors and graft loss. Although a similar protective effect was seen for the CPB2 polymorphism in living donor, this was not significant. Donor kidneys from deceased organ donors have inferior outcomes and lower graft survival rates than kidneys retrieved from living donors.[58] These discrepancies are attributed to the immunological activation, systemic inflammation, and coagulopathies seen in deceased organ donors.[6,27,42] It is therefore not surprising that the protective effect of CPB2 may be stronger in deceased organ donors, especially since C5a levels are higher in these donors compared to their living counterparts.[54,59] Our findings also suggest that donor pre-treatment with CPB2 might be a promising strategy to increase graft function and survival. Recently, a recombinant homolog of CPB2 was shown to alleviate vascular cell damage by decreasing C3a- and C5a-induced neutrophil extracellular trap formation and suggested as an early intervention for COVID-19.[60] Conversely, donor pre-treatment with CBP2 would not be possible until it is known whether other donor organs (i.e., the liver, heart, and lungs) would also benefit from this. Alternatively, rather than enhancing CPB2 levels, blocking the generation of C3a and C5a through a complement inhibitor could be a better approach.[55–57] Lastly, our findings stress the potential importance of early complement inhibition as a potential mitigation factor, as well as that the inhibition needs to be able to get into the kidney allograft given the importance of donor genotypes.

Several limitations of this study warrant consideration. First and foremost, although the associations found are expected to be causal, this cannot be proven by our study because it is observational in nature. Further studies will therefore be needed to confirm that the observed associations are indeed causal. Second, the relationship between polymorphisms and systemic levels of CPB2 could not be determined in our cohort because of a lack of samples, as well as the relationship of CPB2 levels with C3a and C5a concentrations. However, it is important to note, that current available assays can not differentiate between the anaphylatoxin (C3a or C5a) and its metabolite (C3adesArg or C5adesArg). In contrast, crucial strengths of our study are the analysis of four functional polymorphisms in a large sample size of donor-recipient pairs, the clinically meaningful endpoint, and the 15-year follow-up time.

In conclusion, we found that patients receiving a donor kidney carrying the A-allele of the *CPB2* polymorphism rs3742264 have a lower risk of late graft loss. Considering the A-allele is a high-producing *CPB2* variant, our findings imply a beneficial effect of *CPB2* on long-term allograft survival in renal transplantation.

## Disclosure

FP owns stock in Appelis, Iveric Bio, InflaRx and Omeros. AFA is a consultant for Ionis Pharmaceuticals and CSL Behring. JMT and VMH are consultants for Q32 Bio, Inc., a company developing complement inhibitors. Both also hold stock and may receive royalty income from Q32 Bio, Inc. BPD has served as a consultant for Apellis Pharmaceuticals, Alexion AstraZeneca Rare Disease, Novartis Pharmaceuticals, and Arrowhead Pharmaceuticals. The remaining authors of this paper declare that they have no competing interests.

## Data Availability

Anonymized data produced in the present study are available upon reasonable request to the authors.

## Acknowledgment

The authors thank the members of the REGaTTA cohort (REnal GeneTics TrAnsplantation; University Medical Center Groningen, University of Groningen, Groningen, the Netherlands): S. J. L. Bakker, J. van den Born, M. H. de Borst, H. van Goor, J. L. Hillebrands, B. G. Hepkema, G. J. Navis and H. Snieder. The illustrations of Figure 1 were made by Siawosh K. Eskandari.

## Abbreviations

*C3*: Complement protein C3 gene
*C5*: Complement protein C5 gene
CPB2: Carboxypeptidase B2
*CPB2*: Carboxypeptidase B2 gene
*Cpb2*^-/-^: Genetic deficiency of CPB2 in mice
CPN: Carboxypeptidase N
*Cpn*^-/-^: Genetic deficiency of CPN in mice
C statistic: Concordance statistic
C-terminus: Carboxy-terminus
HLA: Human leukocyte antigen
HR: Hazard ratio
IDI: Integrated discrimination improvement
proCPB2: Procarboxypeptidase B2
SNP: Single-nucleotide polymorphism
TAFI: Thrombin-activatable fibrinolysis inhibitor

## Supplementary Data

**Table S1:** Genotypic frequencies of *CPB2* polymorphisms in donors and recipients

| *CPB2* |  | Donor | Recipient | *P*-value<sup>a</sup> | 1000 genome project | *P*-value<sup>b</sup> | *P*-value<sup>c</sup> |
| --- | --- | --- | --- | --- | --- | --- | --- |
| <u>rs2146881</u> | GG | 54.8 (680) | 96.2 (1221) | **<0.0001** | 57.1 (287) | 0.62 | **<0.0001** |
|  | GA | 38.5 (478) | 3.8 (48) |  | 37.2 (187) |  |  |
|  | AA | 6.7 (83) | 0 (0) |  | 5.8 (29) |  |  |
| <u>rs3742264</u> | CC | 45.5 (577) | 44.9 (571) | 0.69 | 44.3 (223) | 0.37 | 0.75 |
|  | CT | 45.2 (574) | 44.8 (569) |  | 44.1 (222) |  |  |
|  | TT | 9.3 (118) | 10.3 (131) |  | 11.5 (58) |  |  |
| <u>rs1926447</u> | GG | 48.6 (617) | 48.8 (618) | 0.65 | 48.1 (242) | 0.60 | 0.22 |
|  | GA | 42.4 (538) | 43.2 (547) |  | 41.4 (208) |  |  |
|  | AA | 9.0 (114) | 8.0 (101) |  | 10.5 (53) |  |  |
| <u>rs3818477</u> | AA | 36.2 (459) | 38.8 (493) | 0.39 | 41.0 (206) | **0.02** | 0.10 |
|  | AC | 50.2 (637) | 47.8 (608) |  | 42.7 (215) |  |  |
|  | CC | 13.6 (172) | 13.4 (170) |  | 16.3 (82) |  |  |

1271 donor-recipient renal transplant pairs were analyzed for the presence of genetic variants in the carboxypeptidase B2 gene (*CPB2*). The frequencies of these polymorphisms were compared to those reported by the European cohort of the 1000 genomes project. Genotype frequencies are displayed as percentages with the corresponding total number of patients [% (n)].
<sup>a</sup> *P*-value for the Pearson Chi-square test for differences in the genotype frequency between donors and recipients.
<sup>b</sup> *P*-value for the Pearson Chi-square test for differences in the genotype frequency between donors and the European cohort of the 1000 genome project.
<sup>c</sup> *P*-value for the Pearson Chi-square test for differences in the genotype frequency between recipients and the European cohort of the 1000 genome project.

**Table S2:**
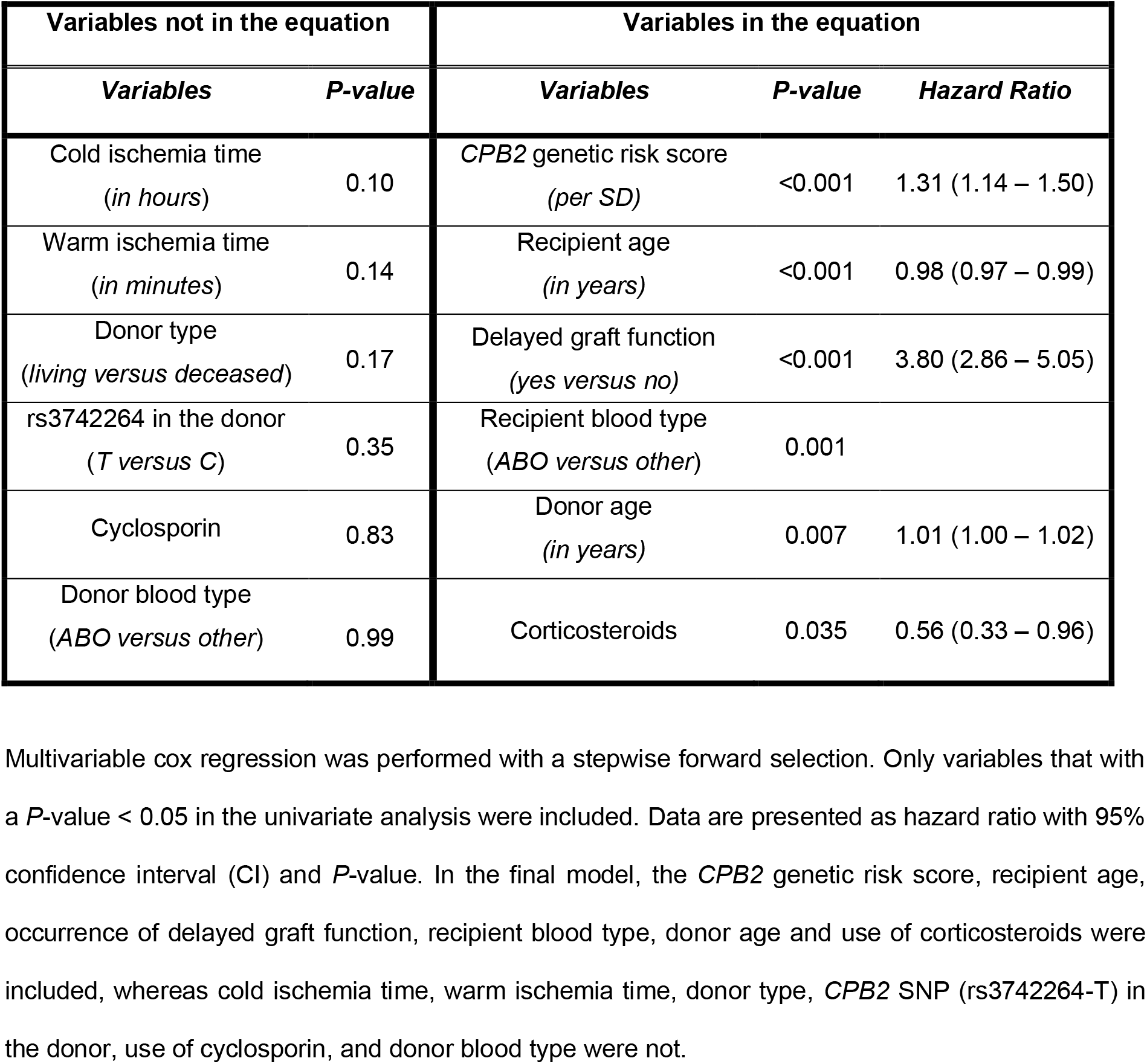
Competitive analysis of clinical factor associations with graft loss.

## Notes

### Funding Statement

This study did not receive any funding.

### Author Declarations

Ethical approval for this study and the study protocol was given by the Institutional Review Board of the University Medical Center Groningen in Groningen, The Netherlands (Medical Ethical Committee 2014/077). The study protocol adhered to the Declaration of Helsinki. All subjects provided written informed consent.

